# Democratic governance and excess mortality during the COVID-19 pandemic

**DOI:** 10.1101/2021.08.25.21262614

**Authors:** Vageesh Jain, Jonathan Clarke, Thomas Beaney

**Affiliations:** Institute for Global Health, University College London, WC1N 1EH, UK; Centre for Mathematics of Precision Healthcare, Department of Mathematics, Imperial College London, SW7 2AZ, UK; Department of Primary Care and Public Health, Imperial College London, W6 8RP, UK

## Abstract

**Background:** Excess mortality has been used to assess the health impact of COVID-19 across countries. Democracies aim to build trust in government and enable checks and balances on decision-making, which may be useful in a pandemic. On the other hand, democratic governments have been criticised as slow to enforce restrictive policies and being overly influenced by public opinion. This study sought to understand whether strength of democratic governance is associated with the variation in excess mortality observed across countries during the pandemic.

**Methods:** Through linking open-access datasets we constructed univariable and multivariable linear regression models investigating the association between country EIU Democracy Index (representing strength of democratic governance on a scale of 0 to 10) and excess mortality rates, from February 2020 to May 2021. We stratified our analysis into high-income and low and middle-income country groups and adjusted for several important confounders.

**Results:** Across 78 countries, the mean EIU democracy index was 6.74 (range 1.94 to 9.81) and the mean excess mortality rate was 128 per 100,000 (range -55 to 503 per 100,000). A one-point increase in EIU Democracy Index was associated with a decrease in excess mortality of 26.3 per 100,000 (p=0.002), after accounting for COVID-19 cases, age ≥ 65, gender, prevalence of cardiovascular disease, universal health coverage and the strength of early government restrictions. This association was particularly strong in high-income countries (β -47.5, p<0.001) but non-significant in low and middle-income countries (β -10.8, p=0.40).

**Conclusions:** Socio-political factors related to the way societies are governed have played an important role in mitigating the overall health impact of COVID-19. Given the omission of such considerations from outbreak risk assessment tools, and their particular significance in high-income countries rated most highly by such tools, this study strengthens the case to broaden the scope of traditional pandemic risk assessment.

**Key Messages:** *What is already known?:*

- Previous studies have found that as countries become more democratic they experience a decline in rates of infant and child mortality, infections such as tuberculosis, and non-communicable diseases.
- In Europe, more democratic countries were initially more reluctant to adopt restrictive COVID-19 measures that could conflict with democratic principles, including lockdowns.

*What are the new findings?:*

- We found that a one-point increase in EIU Democracy Index was associated with a decrease in excess mortality of 26.3 per 100,000 (p=0.002), after accounting for several confounders including demographics, numbers of cases and the strength of early government responses.
- This association was particularly significant in high-income countries (β - 47.5, p<0.001), suggesting that way societies are governed has played an important role in mitigating the impact of COVID-19.

*What do the new findings imply?:*

- Given the omission of social, political and cultural considerations from outbreak risk assessment tools, and criticisms of such tools that have failed to accurately reflect the observed impact of the pandemic across high-income countries, this study builds on the case to broaden of the scope of traditional pandemic risk assessment.
- Future research into the mechanisms underlying our findings will help to understand and address the complex and deep-rooted vulnerabilities countries face in a protracted and large-scale public health emergency.

## Introduction

Excess mortality is widely used as the gold standard in measuring the health impact of COVID-19 across the world [1]. With health systems, services and individual behaviour greatly affected by the pandemic, excess mortality provides an aggregate measure through which to consider both direct COVID-19 and indirect non-COVID-19 deaths due to the pandemic.

Many factors are involved in explaining the variation in excess mortality seen across countries. Studies have clearly demonstrated the importance of pre-existing population-level factors such as increasing age, male gender, comorbidities and obesity in increasing risk of death from COVID-19 [2,3]. Government policy responses to epidemics, including severe restrictions on movement, have also been critical predictors of disease control and deaths [1,4,5]. The drastic measures taken by countries to restrict movement and impose penalties on non-compliance represent a trade-off between health protection and individual freedoms. In the initial phase of the pandemic, governments in some countries were reluctant to adopt measures that conflicted with democratic principles [6] leading to suggestions that such countries were too slow to react [7]. Nevertheless, lockdowns and travel restrictions were quickly used and enforced in both authoritarian countries like China and democratic ones like Taiwan, South Korea and Japan. Even if government restrictions were initially more strongly enforced in authoritarian countries, over the longer-term, mortality from other causes may have increased due to reduced healthcare-seeking behaviours [8,9], the altered provision of routine healthcare, or a rise in mental health issues and risky lifestyle behaviours [10].

Democracies aim to encourage public trust in government, enable checks and balances on decision-making, and increase community participation and social capital [11,12], all of which may be useful in a pandemic. On the other hand, democratic governance has been criticised as enabling the ‘tyranny of the majority [13],’ when decisions are made in the interests of the majority even if harmful to minorities or in conflict with other objectives. With a strong age-based gradient in individual risk due to COVID-19 [3], the pandemic has brought to the fore the inherent tension in restricting the liberties of the masses to protect those more vulnerable. In a representative democracy, the views of the majority should, by design, influence political decision-making, particularly at times of significant uncertainty.

While governments of all countries are responding to the pandemic, a heated debate rages about which political system, democracy versus authoritarian, is better positioned to respond to the pandemic. This study sought to understand whether strength of democratic governance is associated with the variation in excess mortality observed across countries during the COVID-19 pandemic. This will help policymakers to understand how national systems of governance may have altered the impact of COVID-19, informing future plans for pandemic preparedness and global health security.

## Methods

### Data Sources and Extraction

All data used for this study were open-access and available online. Excess mortality data were obtained from the Economist Excess Deaths Tracker [14] representing data on 78 countries. Data spanned time periods from February 2020 to May 2021, with variation in the number of months for which data were available across different countries. Democratic governance at the national level was measured through the Economist Intelligence Unit (EIU) Democracy Index, with each country scored from 0 (least democratic) to 10 (most democratic). This is composed of 60 indicators contained within five domains measuring electoral process, civil liberties, functioning of government, political participation, and political culture. Each country’s Democracy Index was taken from the 2021 EIU report, reflecting the state of democracy in countries after the pandemic had begun. For each country, data were also extracted on a range of routinely available national-level indicators that could confound the relationship between democratic governance and excess mortality. Table 1 shows those selected, their different sources and summary statistics. Data on cumulative COVID-19 cases were obtained from the COVID-19 Data Repository at Johns Hopkins University [15], for the same period of time as data on excess mortality for each country. The Oxford COVID-19 Government Response Tracker Stringency Index is a score out of 100, [16] combining nine different indicators: school closures; workplace closures; cancellation of public events; restrictions on public gatherings; closures of public transport; stay-at-home requirements; public information campaigns; restrictions on internal movements; and international travel controls. A Stringency Index was obtained for each country at the time of their first, 100^th^ and 1000^th^ identified case. Where dates did not align exactly with these case numbers, the closest date was taken.

**Table 1.** Investigated factors, data sources and summary statistics

| Type | Domain | Variable | Period | Source | Mean (SD) | Range |
| --- | --- | --- | --- | --- | --- | --- |
| **Outcome** | Excess mortality | Excess mortality per 100,000 population | Feb 2020 – May 2021 | Economist Excess Deaths Tracker | 128 (120) | -55 - 503 |
| **Exposure** | Democratic governance | EIU Democracy Index | 2020 | Economist Intelligence Unit | 6.74 (1.93) | 1.94 – 9.81 |
| **Covariates** | Demographic | Population age ≥ 65(% total) | 2019 | World Bank | 13.7 (6.20) | 1.00 – 28.0 |
|  |  | Population density (people per sq. km) | 2018 | World Bank | 241 (920) | 2.04 – 7953.0 |
|  |  | Population female (%) | 2019 | World Bank | 50.8 (1.21) | 24.7 – 54.0 |
| | Economic | GDP per capita (current US \$) | 2019 | World Bank | 24236 (23382) | 870.8 – 114704.6 |
|  | Disease burden | Cumulative COVID-19 cases per 100,000 population | Feb 2020-May 2021 | Johns Hopkins University | 4123 (3429) | 0 – 14214.9 |
|  |  | Prevalence of cardiovascular disease (%) | 2019 | Global Burden of Disease Study | 9.51 (3.74) | 0 – 16.5 |
|  |  | Prevalence of chronic respiratory disease (%) | 2019 | Global Burden of Disease Study | 8.00 (3.38) | 0 – 16.1 |
|  |  | Prevalence of diabetes and kidney diseases (%) | 2019 | Global Burden of Disease Study | 16.3 (4.01) | 0 – 25.4 |
|  |  | Prevalence of neurological disorders (%) | 2019 | Global Burden of Disease Study | 40.8 (6.66) | 0 – 50.9 |
|  |  | Prevalence of cancer (%) | 2019 | Global Burden of Disease Study | 11.8 (5.40) | 0 – 27.0 |
|  | Disease risk factors | Prevalence of adult obesity (BMI>30, %) | 2010-2019 | World Obesity Federation | 20.8 (8.04) | 3.6 – 42.7 |
|  |  | Age-standardized mortality rate attributed to household and ambient air pollution per 100,000 population | 2016 | World Health Statistics 2020 | 42.8 (35.7) | 5.9 – 185.2 |
|  |  | Age standardized prevalence of tobacco smoking among persons aged 15 and older (%) | 2018 | World Health Statistics 2020 | 24.0 (8.54) | 6.9 – 44.7 |
|  | Health system | UHC Service Coverage Index | 2017 | World Bank | 75.5 (7.37) | 59 - 89 |
| | | Current health expenditure per capita (current US \$) | 2018 | World Bank | 7.58 (2.38) | 2.49 – 16.9 |
|  |  | Out-of-pocket expenditure (% current health expenditure) | 2018 | World Bank | 28.2 (14.3) | 5.99 – 68.4 |
|  |  | Physicians per 1000 people | 2014-2018 | World Bank | 29.2 (13.0) | 4.5 – 71.2 |
|  | Strength of restrictive policies | COVID-19 Stringency Index | Time of 1<sup>st</sup> case | Oxford COVID-19 Government Response Tracker | 13.3 (12.9) | 0 – 65.7 |
|  |  | COVID-19 Stringency Index | Time of 100<sup>th</sup> case | Oxford COVID-19 Government Response Tracker | 47.9 (29.4) | 0 – 93.5 |
|  |  | COVID-19 Stringency Index | Time of 1000<sup>th</sup> Case | Oxford COVID-19 Government Response Tracker | 67.5 (23.7) | 11.1 – 100 |

### Statistical Analysis

Data were ≥ 95% complete for all investigated variables except smoking and obesity where data were missing for eight and nine countries respectively. Distributions for each variable were examined using histograms and the relationship with excess mortality examined using scatter plots. A log transformation was applied to data on GDP per capita due to a highly negatively skewed distribution. For the proportion of the population who were female, two outliers were identified (Oman and Qatar, with 34.0% and 24.7% female, respectively), but retained for analysis. Univariable regression was performed to investigate which of the extracted variables demonstrated a significant relationship with excess mortality (Table 2). To check for possible multicollinearity [17] between variables, Pairwise Pearson correlation coefficients were calculated (Supplementary File) and although highly collinear both age≥ 65 and prevalence of cardiovascular disease (CVD) were included as they were both considered important predictors of excess mortality from a theoretical point of view. A multivariable linear regression model was constructed to investigate the association between EIU democracy index and excess mortality across countries, controlling for cumulative COVID-19 cases, age ≥ 65, gender, prevalence of CVD, Universal Health Coverage (UHC) Service Index and COVID-19 Stringency Index at the time of the 1000^th^ case in a country. These variables were chosen based through balancing various factors: univariable regression results, multicollinearity (see Supplementary File), data availability (with some variables such as smoking and obesity having more missing data), and theoretical importance, as assessed by study authors. Scatter plots of residuals against fitted values were investigated and showed no violations of heteroskedasticity and quantile plots of residuals showed no departures from normality. Two further multivariable models were constructed to stratify into country income-groups, as defined by the World Bank, of low- and middle-income countries (LMICs) and high-income countries (HICs). Fewer variables were adjusted for in these models to ensure model fit was maintained with smaller numbers. Sensitivity analyses involved constructing further multivariable models to 1) exclude any significant outliers in the high-income country model (Oman/Qatar) and 2) additionally control for a) Stringency Index and b) UHC Index and c) prevalence of CVD, in both of the stratified models (Supplementary File).

**Table 2.** Univariable linear regression of factors associated with excess mortality for all countries

| Variable | Coefficient | P-Value | 95% CI |
| --- | --- | --- | --- |
| EIU Democracy Index | -11.7 | 0.10 | -25.8 – 2.39 |
| Age ≥ 65 years (%) | 2.45 | 0.28 | -1.99 – 6.88 |
| Population density (ppl/sqm) | -0.02 | 0.13 | -0.05 – 0.01 |
| Population female (%) | 33.44 | 0.003 | 11.4 – 55.5 |
| Log GDP per capita (current US \$) | -21.5 | 0.083 | -45.8 – 2.84 |
| Cumulative COVID-19 cases | 0.02 | <0.001 | 0.01 – 0.02 |
| Prevalence of cardiovascular disease (%) | 6.00 | 0.10 | -1.18 – 13.2 |
| Prevalence of chronic respiratory disease (%) | -3.59 | 0.38 | -11.6 – 4.46 |
| Prevalence of diabetes and kidney diseases (%) | 2.37 | 0.49 | -4.44 – 9.18 |
| Prevalence of neurological disorders (%) | -1.73 | 0.40 | -5.83 – 2.36 |
| Prevalence of cancer (%) | 2.23 | 0.38 | -2.82 – 7.27 |
| Prevalence of adult obesity (BMI>30, %) | 0.64 | 0.74 | -3.12 – 4.41 |
| Age-standardized mortality rate attributed to household and ambient air pollution per 100,000 population | 0.34 | 0.38 | -0.43 – 1.11 |
| Age standardized prevalence of tobacco smoking among persons aged 15 and older (%) | 4.03 | 0.02 | 0.78 – 7.27 |
| UHC Service Coverage Index | -3.14 | 0.09 | -6.84 – 0.55 |
| Current health expenditure per capita (% total health expenditure) | 0.10 | 0.99 | -11.55 – 11.74 |
| Out-of-pocket expenditure (% current health expenditure) | 0.81 | 0.41 | -1.12 – 2.74 |
| Number of doctors per 10,000 people | 0.93 | 0.38 | -1.18 – 3.05 |
| COVID-19 Stringency Index at first case nationally | -0.06 | 0.96 | -2.23 – 2.12 |
| COVID-19 Stringency Index at 100<sup>th</sup> case nationally | 1.05 | 0.03 | 0.13 – 1.97 |
| COVID-19 Stringency Index at 1000<sup>th</sup> case nationally | 1.34 | 0.02 | 0.20 – 2.47 |

## Results

A total of 78 countries with available data on excess mortality were included in the analysis: one low-income, 8 lower-middle income, 26 upper middle-income, and 43 high-income countries (as classified by GDP per capita) (Figure 1, panel A). The mean excess mortality across all countries was 128 per 100,000, ranging from -55 to 503 per 100,000 (Figure 1, panel B). A negative excess mortality, meaning fewer deaths occurred than expected based on the pre-pandemic trend, was observed in 13 countries (16.7%). The median number of months for which excess mortality data were available across countries was 11 (IQR 9 – 12.8). The mean EIU democracy index was 6.74, with seven full democracies (scoring 8.01-10), 51 flawed democracies (scoring 6.01-8), 9 hybrid regimes (scoring 4.01-6) and 11 authoritarian regimes (scoring 0 to 4) (Figure 1, panel C).

**Figure 1.**
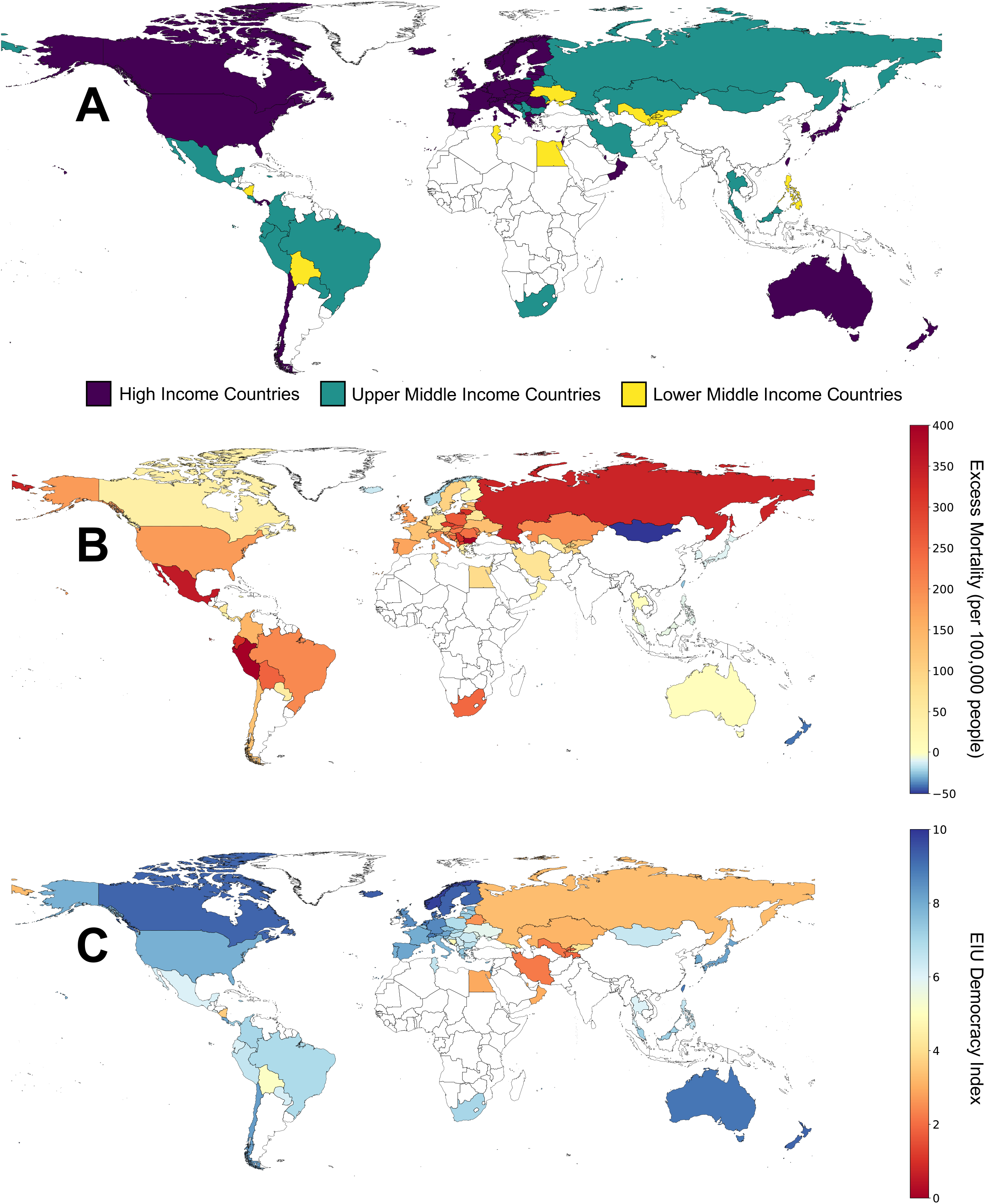
Maps showing the World Bank income classification (panel A), excess mortality per 100,000 people (panel B) and EIU Democracy Index (panel C) of included countries

Table 2 shows the univariable linear regression models for all included variables and excess mortality. The proportion of the population that was female, cumulative COVID-19 cases, prevalence of smoking and COVID-19 Stringency Index at the time of the 100th and 1000th case in countries, were all statistically significantly associated with an increase in excess mortality. Table 3 shows the findings of the multivariable linear regression model. A one-point increase in EIU Democracy Index (scored from 0 to 10) was associated with a statistically significant decrease in excess mortality of 26.3 per 100,000 (p=0.002), after adjusting for cumulative COVID-19 cases, age ≥ 65, gender, prevalence of cardiovascular disease (CVD), Universal Health Coverage (UHC) Service Index and COVID-19 Stringency Index at the time of the 1000th case in a country. A single extra COVID-19 case per 100,000 translated into an increase in excess mortality of 0.2 per 100,000 (p<0.001). A one percent increase in the proportion of the female population was associated with an increase in excess mortality of 9.51 per 100,000. The R2 value for the multivariable model was 0.46.

**Table 3.**
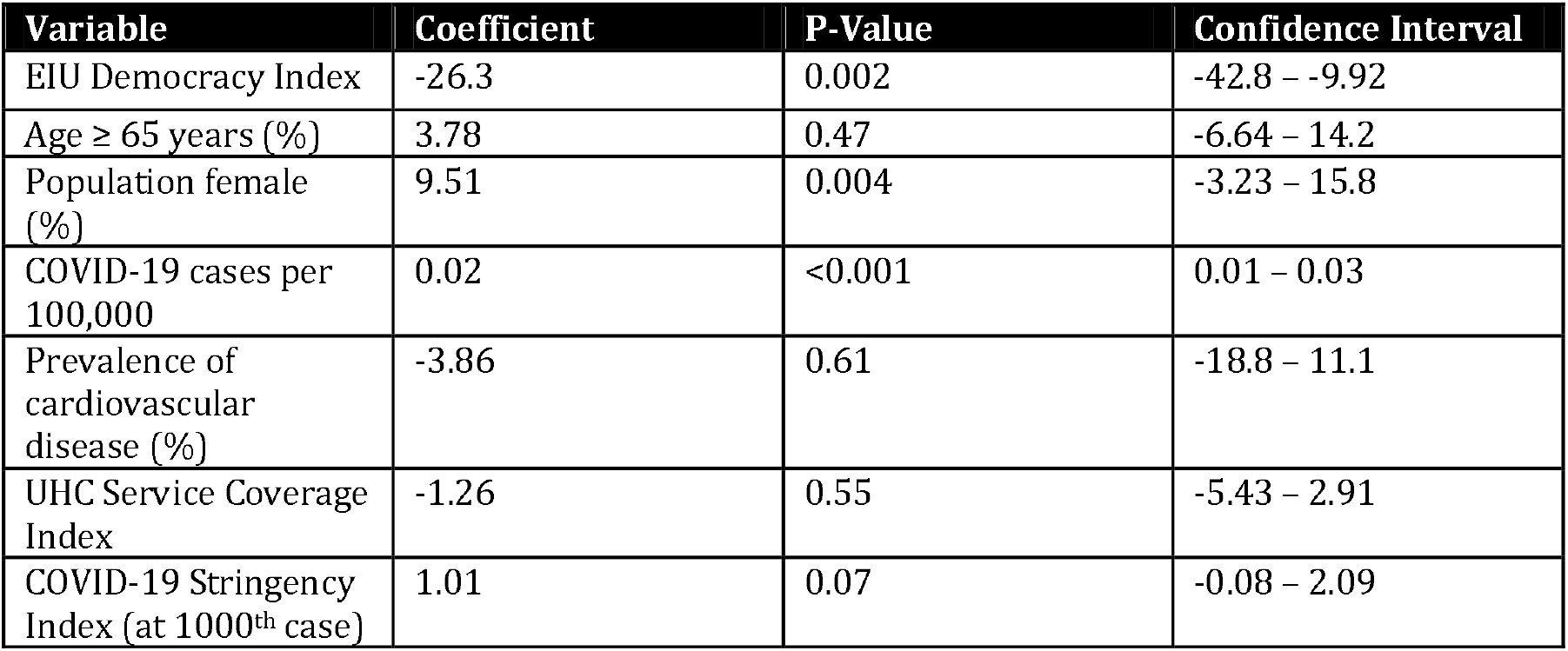
Association between EIU democracy index and excess mortality per 100,000 population

Table 4 shows two further multivariable linear regression models stratified by country income-group. After adjusting for age ≥ 65, gender, and cumulative COVID-19 cases per 100,000 (broadly representing the size of an epidemic), the EIU Democracy Index was not significantly associated with excess mortality in LMICs, but was in HICs, with a one-point increase in the index associated with a decrease in excess mortality of 47.5 per 100,000 (p<0.001). Figure 2 shows the relationship between EIU Democracy Index and excess mortality, with no significant association in LMICs but a linear negative association in HICs. Sensitivity analyses found that 1) the exclusion of two outliers with a low EIU democracy index and a low excess mortality (Oman and Qatar) from the HIC model gave similar results and 2) adding Stringency Index (at the time of either 1st, 100th or 1000th case), UHC index or the prevalence of CVD did not significantly change the strength or magnitude of the estimates across either country income-group model (Supplementary File).

**Table 4.** Association between EIU democracy index and excess mortality per 100,000 population, stratified by country income-group

|  | LMIC (n=34) |  |  | HIC (n=42) |  |  |
| --- | --- | --- | --- | --- | --- | --- |
| Variable | Coefficient | P-Value | Confidence Interval | Coefficient | P-Value | Confidence Interval |
| EIU Democracy Index | -10.8 | 0.40 | -36.5 – 14.9 | -47.5 | <0.001 | -65.4 – -29.5 |
| Age ≥ 65 years (%) | 2.42 | 0.69 | -9.85 – 14.7 | 3.13 | 0.26 | -2.42 – 8.69 |
| Population female (%) | 6.50 | 0.74 | -33.1 – 46.1 | 11.1 | 0.001 | 4.74 – 17.4 |
| COVID-19 cases per 100,000 | 0.03 | 0.004 | 0.01 – 0.05 | 0.02 | <0.001 | 0.01 – 0.02 |

R<sup>2</sup> LMIC model = 0.35, R<sup>2</sup> HIC model = 0.71

**Figure 2.**
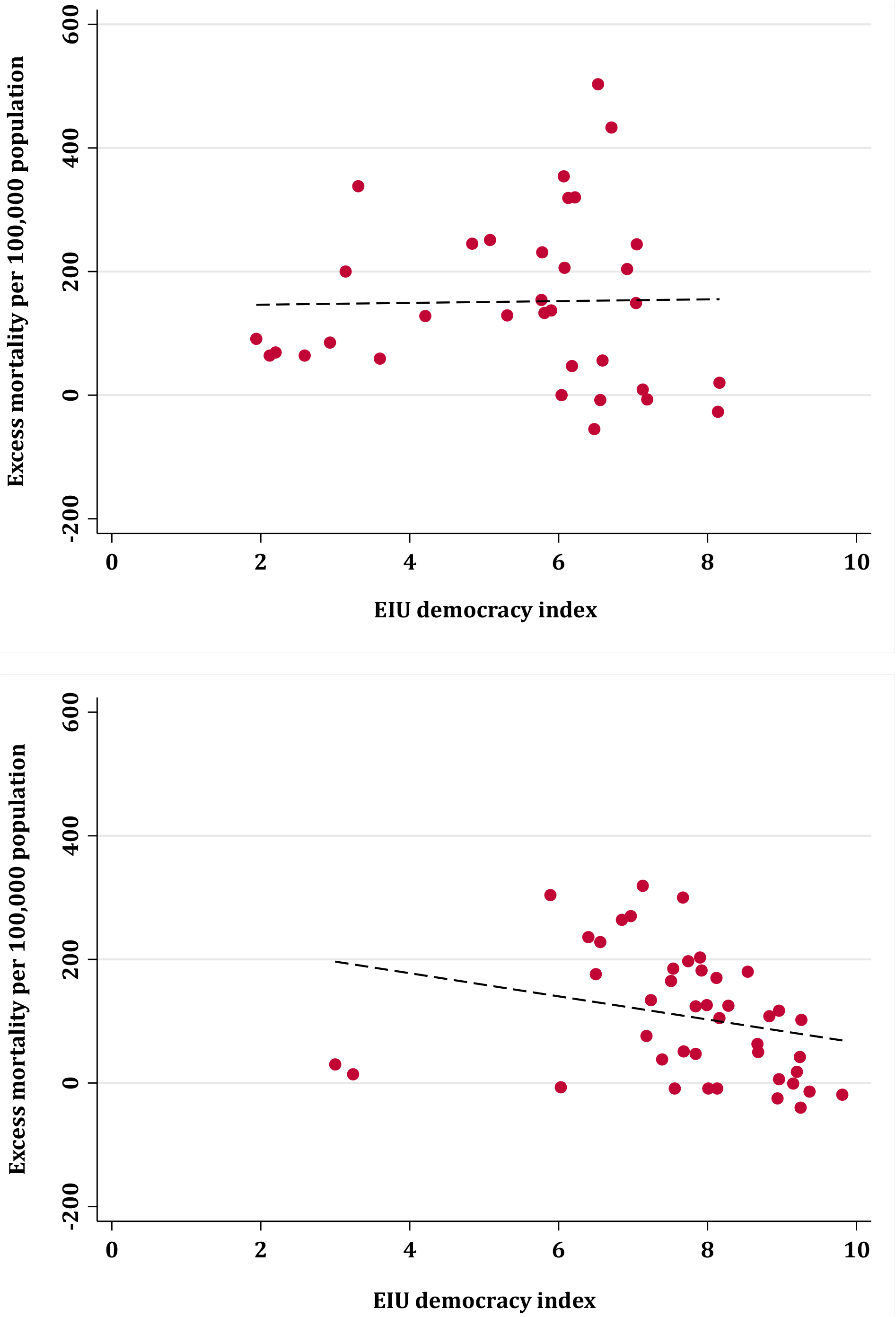
Excess mortality per 100,000 population and EIU Democracy Index across low and middle-income countries (top) and high-income countries (bottom); with line of best fit (dashed)

## Discussion

### Key findings

Nations with higher democratic index scores were associated with having lower rates of excess mortality. This relationship was significant for high-income countries, with results persisting after adjustment for key covariates including the number of recorded COVID-19 cases (i.e. the size of epidemics) and the strength of early government policies to restrict movement, but not low and middle-income countries. This suggests that despite much focus on the timing and severity of reactive policy responses, pre-existing socio-political factors have played an important role in mitigating the overall health impact of COVID-19.

### Democracy and disease control

Democratic principles, such as civil and political liberties, representation, universal suffrage, and free, regular, and fair elections produce competition for popular support among politicians. As such, democracy in theory supports health by ensuring accountability for decisions and actions, and focusing attention on social, economic, and healthcare inequalities [18]. It has been previously observed that as countries become more democratic, they see reductions in mortality across a range of diseases. An ecological study using a range of empirical methods to investigate the relationship between democratic experience and health [19] found that a one-point increase in democratic experience reduced deaths by roughly 2% from cardiovascular diseases, tuberculosis, transport injuries, and other non-communicable diseases combined. Similar studies have concluded that democratic governance has also led to reductions in infant and child mortality [20-22].

There may be various mechanisms underlying this, including government accountability and transparency, the dispersion of power, community participation, social capital, public trust, inclusive and fair institutions, the sharing of data, media freedoms and well-established processes to appraise and implement public health policies [6,18,19,23]. Many of these factors have been vital in the response to COVID-19, affecting public acceptance of and compliance with restrictions [11,12]. Evidence from the Organization for Economic Co-operation and Development (OECD) shows that government’s values, such as high levels of integrity, fairness and openness of institutions are strong predictors of public trust [24]. Trust is necessary for the success of a wide range of public policies that require behavioural responses from the public, including awareness of and compliance with epidemic control policies and guidance on interventions like testing, vaccination and treatment.

We found that even after accounting for COVID-19 case rates (obtained for the same period as excess mortality), more democratic high-income countries had lower rates of excess mortality. This suggests that the mechanisms through which democracy protects against excess deaths extend beyond those that reduce transmission alone. A World Health Organization (WHO) survey found that routine and emergency health services were widely disrupted across countries early on in the pandemic [25]. Observational studies have found large falls in emergency department (ED) attendances in several countries during periods of COVID-19 restrictions [26-28]. A previous study using an instrumented difference-in-difference design found that in England a decline of 2750 ED visits per week for suspected cardiac disease (representing a 35% decrease on pre-pandemic levels), was causally associated with an 18% increase in non-COVID-19 cardiac deaths [28]. In many strongly democratic countries, regular news conferences and press briefings provided reliable, accurate public health information from trusted institutions and experts [18], including on COVID-19 symptoms, testing, and when and how to seek care. The health systems of these countries may also have been better prepared to cope with prolonged periods of disruption. They may have benefited from the ability to provide healthcare remotely [29], strong and flexible leadership and management structures, surge capacity, emergency planning, and well-trained staff committed to quality improvement and evidence-based care [30]. Countries with strongly democratic institutions may have also benefited from large welfare programmes and well-developed public services [31], which may have limited the harmful mental and physical health impacts of social restrictions during the pandemic.

In low and middle-income countries we found there to be no significant association between democratic governance and excess mortality. Given limitations in testing, surveillance systems and death reporting, excess mortality estimates are less reliable than for HICs. Nevertheless, it may also be the case that LMICs face additional challenges which are more important predictors of excess deaths than democratic governance, such as limited access to affordable healthcare [32], vulnerability due to high rates of undiagnosed and poorly controlled chronic conditions [33], and vast numbers pushed into poverty due to the pandemic [34]. With large informal sectors and many relying on daily wages, COVID-19 restrictions were also less extensive in many LMICs compared to HICs. Even severe restrictions appeared to have less of a dramatic impact on population mobility compared to HICs [35]. This means the impact of restrictions on individual behaviours and healthcare attendance may not have been as severely affected, making democratic governance less relevant in this context.

### Other factors explaining excess mortality

Older people and those with co-morbidities are at a greater risk of severe disease, ICU admission and death from COVID-19 [2,3]. We found that although the proportion of the population aged ≥65 was not significantly associated with increased excess mortality, the proportion who were female was, in both the overall and high-income country models. Given systematic differences between men and women in life expectancy, it is likely that the proportion of the population who were female acted as a proxy for very old age, which is much more strongly related with risk of death from COVID-19 (as well as other conditions) compared with risk in the 65-75 age group [36]. Previous evidence suggests that democratic countries are more likely to promote public health and focus on inequalities [19]. If this were true, the populations of highly democratic countries may also have been healthier prior to the pandemic, having a lower risk of death from COVID-19 as well as other conditions. But our analysis finds that even after adjusting for age, gender (a likely proxy for very old age), and the prevalence of cardiovascular disease (a crude measure of preexisting population health), the significant negative association between democratic governance and excess mortality persists.

Although democratic governance covers a range of important factors related to disease control, there are other important social, cultural and political factors that may not be captured by this indicator. A 2021 cross-country regression analysis [37] found that tight cultures, which have strict norms and punishments for deviance, were better able to respond to COVID-19 compared to loose cultures, which have more permissive norms. Nations with high levels of cultural looseness were estimated to have had five times the number of cases (7132 per million vs 1428 per million, respectively) and almost nine times the number of deaths (183 per million vs 21 per million, respectively) compared to those with high levels of cultural tightness. Similarly, it has been proposed that more collectivist societies, which include many East Asian countries, have performed better than more individualist societies in combating COVID-19 [38]. Hofstede’s dimensions of national culture [39], capturing cultural differences such as individualism versus collectivism, uncertainty avoidance, and power distance, may further help to explain the variation in impact of COVID-19 across countries, although many of these factors may overlap with strength of democratic governance to some extent.

### Strengths and limitations

Unlike previous ecological studies on the socio-political determinants of COVID-19 [6,37,40] we used excess mortality as our outcome measure rather than COVID-19 cases or deaths. This provides a more comprehensive and unbiased assessment of the impact of COVID-19 within countries, given significant international variations in testing capacity and differing practices on reporting causes of death [1]. Through linking routine datasets we were also able to control for a range of important confounders, including the size of epidemics and the strength of government restrictions on movement, when assessing the relationship between excess mortality and democratic governance.

The first major limitation of this study is an inability to draw causal inference. Strength of democratic governance is associated with a range of other factors, such as country income. Although we accounted for this through stratification, and for other factors through adjustment in our multivariable model, due to the observational nature of this study with limited available data, it is not possible to rule out all potential confounders. Still, we believe this analysis provides robust evidence of a significant association between strength of democratic governance and excess mortality at the international level. Second, the EIU Democracy Index is not the only measure of democratic governance available, and may not entirely reflect how political values and practices have changed in countries over the course of the COVID-19 response. Nevertheless, the estimates used cover a wide range of 60 indicators and were published in a 2021 report using information from the entirety of 2020, representing the most up-to-date and robust international score on democratic governance available. Finally, we were not able to obtain excess mortality data for all countries, limiting the generalizability of our findings. Nevertheless, we were still able to include 78 countries in the analysis (and a majority of 42 out of 77 (55%) high-income countries), representing a significant step forward in the literature making use of such routine data to understand variation in the impact of COVID-19 across countries.

### Implications for future research and policy

At the national level, recommendations on improving pandemic response focus on compliance with International Health Regulations through specific public health capacities such as surveillance, testing, communications, and countermeasures [41,42]. Our findings suggest that the way societies are governed can also alter the impact of epidemics. Unlike previous research following countries over time as they became more democratic [19], we cannot conclude from our cross-sectional analysis that strengthening democratic institutions within countries will improve pandemic response. But given the relative omission of socio-political considerations from outbreak risk assessment tools [43], and their apparent significance in high-income countries rated most highly by such tools, this study strengthens the case for further research into expanding the scope of traditional pandemic risk assessment.

Operationalizing our findings in practice will require a better understanding of the underlying mechanisms. It is necessary to understand which particular features of strong democracies are most advantageous during a health emergency. It is also important to evaluate the long-term impact of restrictive disease control legislation in the context of democratic governance. Changes in the law have been widely used during the pandemic, prompting WHO to establish a COVID-19 Law Lab to help nations implement legal frameworks [44]. In many cases legal intervention has been effective in curbing disease transmission. For instance, U.S States where masks were legally mandated had significantly fewer COVID-19 cases than those where they were not [45]. Understanding how aligned such restrictions are with democratic principles, the trade-offs individuals are willing to make and their circumstances, are important questions for future research.

## Conclusions

Strength of democratic governance was associated with lower rates of excess mortality during the COVID-19 pandemic, particularly in high-income countries. Several other factors help to explain international variation in excess mortality, including population demographics, the size of COVID-19 epidemics and the severity of early lockdowns, but these do not counterbalance the effect of democratic governance. Future research is needed to better understand the mechanisms underlying this. Recent pandemic response reviews have proposed a number of specific reforms, with an emphasis on narrow public health capacities and reactive government policies. Our study suggests that considering more long-term and deep-rooted socio-political factors may aid pandemic preparedness risk assessments in better capturing the complex vulnerabilities countries face in a protracted and large-scale public health emergency.

## Data Availability

Data underpinning this analysis is available upon request and will be made widely available and uploaded after the paper has been peer-reviewed

## Authors and Contributors

All authors contributed to all stages of this study, including inception, design, literature search, data extraction, data analysis, data interpretation, illustrations, write-up, editing and revisions.

Substantial contributions to the conception or design of the work; or the acquisition, analysis, or interpretation of data for the work: VJ, JC, TB

Drafting the work or revising it critically for important intellectual content: VJ, JC, TB

Final approval of the version to be published: VJ, JC, TB

Agreement to be accountable for all aspects of the work in ensuring that questions related to the accuracy or integrity of any part of the work are appropriately investigated and resolved: VJ, JC, TB

## Ethics Approval

This study did not receive nor require ethics approval, as it does not involve human & animal participants.

## References

[1] Beaney T, Clarke JM, Jain V, et al. Excess mortality: the gold standard in measuring the impact of COVID-19 worldwide? J R Soc Med 2020;113(9):329–334.

[2] Jain V, Yuan J. Predictive symptoms and comorbidities for severe COVID-19 and intensive care unit admission: a systematic review and meta-analysis. International journal of public health 2020;65:533–546.

[3] Williamson EJ, Walker AJ, Bhaskaran K, et al. Factors associated with COVID-19-related death using OpenSAFELY. Nature 2020;584(7821):430–436.

[4] Han E, Tan MMJ, Turk E, et al. Lessons learnt from easing COVID-19 restrictions: an analysis of countries and regions in Asia Pacific and Europe. The Lancet 2020.

[5] Patel J, Sridhar D. We should learn from the Asia–Pacific responses to COVID-19. The Lancet Regional Health–Western Pacific 2020;5.

[6] Engler S, Brunner P, Loviat R, et al. Democracy in times of the pandemic: explaining the variation of COVID-19 policies across European democracies. West European Politics 2021:1–22.

[7] Sebhatu A, Wennberg K, Arora-Jonsson S, et al. Explaining the homogeneous diffusion of COVID-19 nonpharmaceutical interventions across heterogeneous countries. Proc Natl Acad Sci U S A 2020 Sep 1;117(35):21201–21208.

[8] McIntosh A, Bachmann M, Siedner MJ, et al. Effect of COVID-19 lockdown on hospital admissions and mortality in rural KwaZulu-Natal, South Africa: interrupted time series analysis. BMJ Open 2021 Mar 18;11(3):e047961-2020-047961.

[9] Butt JH, Fosbøl EL, Gerds TA, et al. All-cause mortality and location of death in patients with established cardiovascular disease before, during, and after the COVID-19 lockdown: a Danish Nationwide Cohort Study. Eur Heart J 2021;42(15):1516–1523.

[10] Niedzwiedz CL, Green MJ, Benzeval M, et al. Mental health and health behaviours before and during the initial phase of the COVID-19 lockdown: longitudinal analyses of the UK Household Longitudinal Study. J Epidemiol Community Health 2021 Mar;75(3):224–231.

[11] Wong AS, Kohler JC. Social capital and public health: responding to the COVID-19 pandemic. Globalization and Health 2020;16(1):1–4.

[12] Pak A, McBryde E, Adegboye OA. Does high public trust amplify compliance with stringent COVID-19 government health guidelines? A multi-country analysis using data from 102,627 individuals. Risk Management and Healthcare Policy 2021;14:293.

[13] Horwitz MJ. Tocqueville and the Tyranny of the Majority. The review of politics 1966;28(3):293–307.

[14] The Economist. Tracking covid-19 excess deaths across countries. 2021; Available at: https://www.economist.com/graphic-detail/coronavirus-excess-deaths-tracker. Accessed 03/15, 2021.

[15] Johns Hopkins University. COVID-19 Data Repository by the Center for Systems Science and Engineering (CSSE) at Johns Hopkins University. 2021; Available at: https://github.com/CSSEGISandData/COVID-19. Accessed 07/15, 2021.

[16] Our World in Data. COVID-19: Stringency Index. 2021; Available at: https://ourworldindata.org/covid-stringency-index. Accessed 05/20, 2021.

[17] Dormann CF, Elith J, Bacher S, et al. Collinearity: a review of methods to deal with it and a simulation study evaluating their performance. Ecography 2013;36(1):27–46.

[18] Ruger JP. Social justice as a foundation for democracy and health. BMJ 2020;371.

[19] Bollyky TJ, Templin T, Cohen M, et al. The relationships between democratic experience, adult health, and cause-specific mortality in 170 countries between 1980 and 2016: an observational analysis. The Lancet 2019;393(10181):1628–1640.

[20] Wigley S, Akkoyunlu-Wigley A. The impact of democracy and media freedom on under-5 mortality, 1961–2011. Soc Sci Med 2017;190:237–246.

[21] Kudamatsu M. Has democratization reduced infant mortality in sub-Saharan Africa? Evidence from micro data. Journal of the European Economic Association 2012;10(6):1294–1317.

[22] Pieters H, Curzi D, Olper A, et al. Effect of democratic reforms on child mortality: a synthetic control analysis. The Lancet Global Health 2016;4(9):e627–e632.

[23] Ruger JP. Democracy and health. QJM 2005;98(4):299–304.

[24] Organization for Economic Co-operation and Development. Trust in Government. 2021; Available at: https://www.oecd.org/governance/trust-in-government.htm. Accessed 07/21, 2021.

[25] World Health Organization. COVID-19 significantly impacts health services for noncommunicable diseases. 2020; Available at: https://www.who.int/news-room/detail/01-06-2020-covid-19-significantly-impacts-health-services-for-noncommunicable-diseases. Accessed 06/01, 2020.

[26] Oikonomou E, Aznaouridis K, Barbetseas J, et al. Hospital attendance and admission trends for cardiac diseases during the COVID-19 outbreak and lockdown in Greece. Public Health 2020;187:115–119.

[27] McDonnell T, Nicholson E, Conlon C, et al. Assessing the impact of COVID-19 public health stages on paediatric emergency attendance. International journal of environmental research and public health 2020;17(18):6719.

[28] Katsoulis M, Gomes M, Lai AG, et al. Estimating the effect of reduced attendance at emergency departments for suspected cardiac conditions on cardiac mortality during the COVID-19 pandemic. Circulation: Cardiovascular Quality and Outcomes 2021;14(1):e007085.

[29] BMJ. The “virtual wards” supporting patients with covid-19 in the community. 2020; Available at: https://www.bmj.com/content/369/bmj.m2119. Accessed 06/08, 2020.

[30] Nuzzo JB, Meyer D, Snyder M, et al. What makes health systems resilient against infectious disease outbreaks and natural hazards? Results from a scoping review. BMC Public Health 2019;19(1):1–9.

[31] Borrell C, Espelt A, Rodriguez-Sanz M, et al. Politics and health. J Epidemiol Community Health 2007 Aug;61(8):658–659.

[32] Okereke M, Ukor NA, Adebisi YA, et al. Impact of COVID-19 on access to healthcare in low-and middle-income countries: current evidence and future recommendations. Int J Health Plann Manage 2021;36(1):13–17.

[33] Dunachie S, Chamnan P. The double burden of diabetes and global infection in low and middle-income countries. Trans R Soc Trop Med Hyg 2019;113(2):56–64.

[34] World Bank Group. Updated estimates of the impact of COVID-19 on global poverty: Looking back at 2020 and the outlook for 2021. 2021; Available at: https://blogs.worldbank.org/opendata/updated-estimates-impact-covid-19-global-poverty-looking-back-2020-and-outlook-2021. Accessed 03/30, 2021.

[35] Fakir AM, Bharati T. Pandemic catch-22: The role of mobility restrictions and institutional inequalities in halting the spread of COVID-19. Plos one 2021;16(6):e0253348.

[36] Centers for Disease Control and Prevention (CDC). People Who Are at Higher Risk for Severe Illness. 2020; Available at: https://www.cdc.gov/coronavirus/2019-ncov/need-extra-precautions/people-at-higher-risk.html. Accessed 04/20, 2020.

[37] Gelfand MJ, Jackson JC, Pan X, et al. The relationship between cultural tightness–looseness and COVID-19 cases and deaths: a global analysis. The Lancet Planetary Health 2021;5(3):e135–e144.

[38] Liu JH. Majority world successes and European and American failure to contain COVID-19: Cultural collectivism and global leadership. Asian Journal of Social Psychology 2021;24(1):23–29.

[39] Hofstede G. Dimensionalizing cultures: The Hofstede model in context. Online readings in psychology and culture 2011;2(1):2307-0919.1014.

[40] Arachchi JI, Managi S. The role of social capital in COVID-19 deaths. BMC Public Health 2021;21(1):1–9.

[41] World Health Organization.Review Committee on the Functioning of the International Health Regulations (2005) during the COVID-19 Response. 2020; Available at: https://www.who.int/teams/ihr/ihr-review-committees/covid-19. Accessed 03/30, 2021.

[42] Sirleaf EJ, Clark H. Report of the Independent Panel for Pandemic Preparedness and Response: making COVID-19 the last pandemic. The Lancet 2021.

[43] Boyd MJ, Wilson N, Nelson C. Validation analysis of Global Health Security Index (GHSI) scores 2019. BMJ Glob Health 2020 Oct;5(10):10.1136/bmjgh-2020-003276.

[44] World Health Organization. New COVID-19 Law Lab to provide vital legal information and support for the global COVID-19 response. 2021; Available at: https://www.who.int/news/item/22-07-2020-new-covid-19-law-lab-to-provide-vital-legal-information-and-support-for-the-global-covid-19-response. Accessed 07/22, 2021.

[45] Lyu W, Wehby GL. Community Use Of Face Masks And COVID-19: Evidence From A Natural Experiment Of State Mandates In The US: Study examines impact on COVID-19 growth rates associated with state government mandates requiring face mask use in public. Health Aff 2020;39(8):1419–1425.

