## Supplementary File for "Democratic governance and excess mortality during the COVID-19 pandemic"

**Table 1 – Pearson’s correlation plot for all included variables**

|  | **EIU Index** | **Age ≥ 65** | **Pop density** | **Female** | **GDP per cap** | **COVID cases** | **Prevalence CVD** | **Prevalence respiratory disease** | **Prevalence diabetes/kidney disease** | **Prevalence neurological disease** | **Prevalence Cancer** | **Obesity** | **Smoking** | **Pollution** | **UHC** | **% GDP on health** | **Out-of-pocket expenditure** | **Physicians per 1000 population** | **Stringency Index at 1^st^ case** | **Stringency Index at 100^th^ case** | **Stringency Index at 1000^th^ case** |
| --- | --- | --- | --- | --- | --- | --- | --- | --- | --- | --- | --- | --- | --- | --- | --- | --- | --- | --- | --- | --- | --- |
| **EIU Index** | 1.00 |  |  |  |  |  |  |  |  |  |  |  |  |  |  |  |  |  |  |  |  |
| **Age** | 0.61 | 1.00 |  |  |  |  |  |  |  |  |  |  |  |  |  |  |  |  |  |  |  |
| **Pop density** | 0.00 | -0.01 | 1.00 |  |  |  |  |  |  |  |  |  |  |  |  |  |  |  |  |  |  |
| **Female** | -0.12 | 0.35 | -0.33 | 1.00 |  |  |  |  |  |  |  |  |  |  |  |  |  |  |  |  |  |
| **GDP** | 0.59 | 0.42 | 0.23 | -0.27 | 1.00 |  |  |  |  |  |  |  |  |  |  |  |  |  |  |  |  |
| **COVID cases** | 0.30 | 0.47 | -0.10 | 0.20 | 0.29 | 1.00 |  |  |  |  |  |  |  |  |  |  |  |  |  |  |  |
| **Prevalence CVD** | 0.37 | 0.88 | -0.08 | 0.46 | 0.26 | 0.46 | 1.00 |  |  |  |  |  |  |  |  |  |  |  |  |  |  |
| **Prevalence respiratory disease** | 0.66 | 0.65 | -0.04 | -0.10 | 0.67 | 0.40 | 0.45 | 1.00 |  |  |  |  |  |  |  |  |  |  |  |  |  |
| **Prevalence diabetes/kidney disease** | 0.34 | 0.68 | 0.13 | 0.36 | 0.18 | 0.34 | 0.71 | 0.35 | 1.00 |  |  |  |  |  |  |  |  |  |  |  |  |
| **Prevalence neurological disease** | 0.39 | 0.73 | -0.02 | 0.19 | 0.44 | 0.36 | 0.67 | 0.63 | 0.58 | 1.00 |  |  |  |  |  |  |  |  |  |  |  |
| **Prevalence Cancer** | 0.12 | 0.60 | 0.26 | 0.31 | 0.28 | 0.26 | 0.69 | 0.16 | 0.50 | 0.48 | 1.00 |  |  |  |  |  |  |  |  |  |  |
| **Obesity** | -0.30 | -0.37 | -0.21 | 0.13 | -0.13 | -0.06 | -0.23 | -0.10 | -0.16 | -0.21 | -0.29 | 1.00 |  |  |  |  |  |  |  |  |  |
| **Smoking** | 0.08 | 0.46 | -0.10 | 0.34 | -0.11 | 0.33 | 0.54 | 0.12 | 0.36 | 0.37 | 0.41 | -0.14 | 1.00 |  |  |  |  |  |  |  |  |
| **Pollution** | -0.59 | -0.53 | -0.08 | 0.00 | -0.61 | -0.32 | -0.29 | -0.60 | -0.34 | -0.40 | -0.20 | -0.03 | 0.14 | 1.00 |  |  |  |  |  |  |  |
| **UHC** | 0.56 | 0.44 | 0.20 | -0.12 | 0.69 | 0.19 | 0.18 | 0.66 | 0.14 | 0.45 | 0.15 | -0.13 | -0.27 | -0.73 | 1.00 |  |  |  |  |  |  |
| **% GDP on health** | 0.54 | 0.54 | -0.13 | -0.06 | 0.43 | 0.30 | 0.39 | 0.70 | 0.20 | 0.45 | 0.14 | 0.00 | 0.15 | -0.49 | 0.48 | 1.00 |  |  |  |  |  |
| **Out-of-pocket expenditure** | -0.51 | -0.37 | 0.02 | -0.03 | -0.55 | -0.36 | -0.24 | -0.49 | -0.16 | -0.25 | -0.16 | 0.08 | 0.15 | 0.64 | -0.54 | -0.34 | 1.00 |  |  |  |  |
| **Physicians per 1000 population** | 0.28 | 0.61 | -0.06 | 0.45 | 0.32 | 0.44 | 0.60 | 0.35 | 0.41 | 0.57 | 0.43 | -0.03 | 0.34 | -0.31 | 0.27 | 0.22 | -0.17 | 1.00 |  |  |  |
| **Stringency Index at 1^st^ case** | -0.28 | -0.36 | 0.09 | 0.01 | -0.27 | -0.20 | -0.31 | -0.29 | -0.34 | -0.28 | -0.19 | -0.09 | -0.06 | 0.46 | -0.28 | -0.28 | 0.24 | -0.08 | 1.00 |  |  |
| **Stringency Index at 100^th^ case** | -0.28 | -0.26 | -0.07 | 0.17 | -0.46 | -0.08 | -0.14 | -0.42 | -0.12 | -0.36 | -0.15 | 0.04 | 0.03 | 0.32 | -0.53 | -0.36 | 0.34 | -0.09 | 0.43 | 1.00 |  |
| **Stringency Index at 1000^th^ case** | -0.23 | -0.30 | -0.19 | 0.15 | -0.41 | 0.00 | -0.18 | -0.39 | -0.22 | -0.35 | -0.08 | 0.11 | -0.03 | 0.34 | -0.49 | -0.49 | 0.18 | -0.16 | 0.29 | 0.63 | 1.00 |

**Table 2 – Association between excess mortality per 100,000 population and democratic governance, excluding high-income country outliers**

|  | HIC (n=40) |  |  |
| --- | --- | --- | --- |
| Variable | **Coefficient** | **P-Value** | **Confidence Interval** |
| EIU Democracy Index | -44.8 | <0.001 | -65.8 - -23.8 |
| Age 65+ years (%) | 1.68 | 0.58 | -4.35 – 7.72 |
| Population female (%) | 20.8 | 0.04 | 0.81 – 40.9 |
| COVID-19 cases per 100,000 | 0.015 | <0.001 | 0.009 – 0.021 |

**Table 3 - Association between excess mortality per 100,000 population and democratic governance, excluding outliers and adding stringency index at 1^st^ case**

|  | LMIC (n=33) |  |  | HIC (n=39) |  |  |
| --- | --- | --- | --- | --- | --- | --- |
| Variable | **Coefficient** | **P-Value** | **Confidence Interval** | **Coefficient** | **P-Value** | **Confidence Interval** |
| EIU Democracy Index | -14.1 | 0.26 | -39.4 – 11.1 | -27.8 | 0.02 | -50.7 - -4.77 |
| Age 65+ years (%) | 2.90 | 0.65 | -10.2 – 16.0 | 1.69 | 0.54 | -3.81 – 7.19 |
| Population female (%) | 0.91 | 0.96 | -40.2 – 42.0 | 28.0 | 0.005 | 8.92 – 47.0 |
| COVID-19 cases per 100,000 | 0.04 | 0.001 | 0.02 – 0.05 | 0.02 | <0.001 | 0.01 – 0.02 |
| Stringency Index at 1^st^ case | -0.20 | 0.88 | -2.84 – 2.43 | 2.04 | 0.19 | -1.06 – 5.14 |

**Table 4 - Association between excess mortality per 100,000 population and democratic governance, excluding outliers and adding stringency index at 1^st^ case**

|  | LMIC (n=33) |  |  | HIC (n=39) |  |  |
| --- | --- | --- | --- | --- | --- | --- |
| Variable | **Coefficient** | **P-Value** | **Confidence Interval** | **Coefficient** | **P-Value** | **Confidence Interval** |
| EIU Democracy Index | -14.7 | 0.25 | -40.3 – 10.9 | -37.4 | 0.002 | -60.2 - -14.6 |
| Age 65+ years (%) | 3.28 | 0.58 | -8.68 – 15.3 | 1.34 | 0.64 | -4.49 – 7.17 |
| Population female (%) | -0.09 | 0.99 | -39.2 – 39.0 | 27.4 | 0.01 | 7.07 – 47.8 |
| COVID-19 cases per 100,000 | 0.03 | 0.001 | 0.01 – 0.05 | 0.02 | <0.001 | 0.01 – 0.02 |
| Stringency Index at 100^th^ case | 0.14 | 0.84 | -1.27 – 1.55 | -0.32 | 0.52 | -1.31 – 0.67 |

**Table 5 - Association between excess mortality per 100,000 population and democratic governance, excluding outliers and adding stringency index at 1^st^ case**

|  | LMIC (n=33) |  |  | HIC (n=39) |  |  |
| --- | --- | --- | --- | --- | --- | --- |
| Variable | **Coefficient** | **P-Value** | **Confidence Interval** | **Coefficient** | **P-Value** | **Confidence Interval** |
| EIU Democracy Index | -20.7 | 0.12 | -46.9 – 5.58 | -36.5 | 0.001 | -57.6 - -15.4 |
| Age 65+ years (%) | 4.59 | 0.43 | -7.14 – 16.3 | 1.35 | 0.63 | -4.27 – 6.97 |
| Population female (%) | -0.83 | 0.96 | -38.7 – 37.0 | 28.4 | 0.006 | 8.69 – 48.1 |
| COVID-19 cases per 100,000 | 0.03 | 0.002 | 0.01 – 0.05 | 0.02 | <0.001 | 0.01 – 0.02 |
| Stringency Index at 1000^th^ case | 1.43 | 0.18 | -0.71 – 3.58 | -0.47 | 0.28 | -1.33 – 0.40 |

**Table 6 - Association between excess mortality per 100,000 population and democratic governance, excluding outliers and adding universal health coverage service index**

|  | LMIC (n=34) |  |  | HIC (n=40) |  |  |
| --- | --- | --- | --- | --- | --- | --- |
| Variable | **Coefficient** | **P-Value** | **Confidence Interval** | **Coefficient** | **P-Value** | **Confidence Interval** |
| EIU Democracy Index | -10.7 | 0.40 | -36.4 – 15.1 | -44.7 | 0.001 | -70.8 - -18.5 |
| Age 65+ years (%) | 5.01 | 0.45 | -8.42 – 18.4 | 1.69 | 0.58 | -4.47 – 7.85 |
| Population female (%) | -1.37 | 0.95 | -44.3 – 41.6 | 20.7 | 0.08 | -2.62 – 44.1 |
| COVID-19 cases per 100,000 | 0.03 | 0.008 | 0.01 – 0.04 | 0.02 | <0.001 | 0.01 – 0.02 |
| UHC Service Coverage Index | 4.01 | 0.33 | -4.33 – 12.3 | -0.05 | 0.99 | -5.65 – 5.55 |

**Table 7 - Association between excess mortality per 100,000 population and democratic governance, excluding outliers and adding prevalence of CVD**

|  | LMIC (n=34) |  |  | HIC (n=40) |  |  |
| --- | --- | --- | --- | --- | --- | --- |
| Variable | **Coefficient** | **P-Value** | **Confidence Interval** | **Coefficient** | **P-Value** | **Confidence Interval** |
| EIU Democracy Index | -21.7 | 0.13 | -49.8 – 6.41 | -41.8 | <0.001 | -62.6 - -21.0 |
| Age 65+ years (%) | 22.3 | 0.10 | -4.20 – 48.8 | -3.04 | 0.46 | -11.2– 5.15 |
| Population female (%) | 5.30 | 0.78 | -33.1 – 43.8 | 16.6 | 0.11 | -3.67 – 36.8 |
| COVID-19 cases per 100,000 | 0.03 | 0.007 | 0.01 – 0.04 | 0.01 | <0.001 | 0.01 – 0.02 |
| Prevalence CVD | -27.0 | 0.10 | -59.1 – 5.17 | 9.75 | 0.10 | -2.01 – 21.5 |
